# Concordance of weight status between mothers and children: A secondary analysis of the Pakistan Demographic and Health Survey VII

**DOI:** 10.1101/2023.05.04.23289509

**Authors:** Faiz Alam, Mohammed K. Ali, Shivani A. Patel, Romaina Iqbal

## Abstract

Familial concordance of nutritional status is an emerging field of study that may guide the development of interventions that operate beyond the individual and within the family context. Little published data exist for concordance of nutritional status within Pakistani households. We assessed the associations between weight status of mothers and their children in a nationally representative sample of households in Pakistan using Demographic and Health Survey data. Our analysis included 3465 mother-child dyads, restricting to children under-five years of age with body mass index (BMI) information on their mothers. We used linear regression models to assess the associations between maternal BMI category (underweight, normal weight, overweight, obese) and child’s weight-for-height z-score (WHZ), accounting for socio-demographic characteristics of mothers and children. We assessed these relationships in all children under-five and also stratified by age of children (younger than 2 years and 2 to 5 years). In all children under-five and in children 2 to 5 years, maternal BMI was positively associated with child’s WHZ, while there was no association between maternal BMI and child WHZ for children under-two. The findings indicate that the weight status of mother’s is positively associated with that of their children. These associations have implications for interventions aimed at healthy weights of families.

## Introduction

There is a growing body of literature focusing on concordance of cardiometabolic disease risk factors and outcomes within families and households, suggesting that NCD prevention programs may be more effective if the family – rather than the individual – was targeted for intervention. Studies show increased likelihood of adults living with someone with a chronic condition, such as obesity, diabetes, hypertension, hyperlipidemia, and other cardiometabolic conditions, having that same condition. This includes concordance between parent-child [1,2], sibling [3,4], spousal dyads [5], and between any co-residing members of households [6,7]. These studies investigate the genetic predisposition to disease [6], as can be seen in strictly familial relationships, and also the effect of the shared environment [8] and health behavior influences [9,10], such as in co-residing studies.

Intergenerational concordance within households – and specifically that between parents and children – may reflect a composite of genetic, epigenetic, and shared environmental factors. The magnitude of intergenerational concordance of NCD risk within households in Pakistan is largely unknown. Studies focusing on parent-child concordance have been primarily done in high-income countries, such as in the United Kingdom and Australia [11–16]. There is some evidence from low- and middle-income countries (LMICs) regarding concordance of biomarkers such as hemoglobin A1c and C-reactive protein [2]. Specifically in South Asia, studies show statistically significant associations between child under-five obesity and maternal weight, but do not further stratify these relationships by child age [17,18].

Within Pakistan, there is particular importance of the parent-child dyad. Pakistan has one of the highest average household sizes and percentage of multigenerational households in the world, suggesting extended sharing of environment between generations. Moreover, Pakistan is following a global trend of increasing life expectancy, signaling a large population of individuals at higher risk of developing NCDs that the Pakistani healthcare system will have to manage in the future [19]. Additionally, children in Pakistan are becoming increasingly vulnerable to different forms of malnutrition: in the 2018 Pakistan National Nutrition Survey (PNNS), wasting was noted in 17.7% of children, the highest in Pakistan’s history, and prevalence of overweight children under-five almost doubled from 5% in 2011 to 9.5% in 2018. Similarly, women ages 15-49 experienced an increase in overweight and obesity increasing from 28% to 37.8% from 2011 to 2018 [20]. This parallel increase in excess weight across a wide range of Pakistani age demographics may have important implications for cardiometabolic disease at the population level. Thus, there is a need to study Pakistani household concordance of NCD risk factors to further understand the extent, if present, of household concordance in Pakistan, especially with an additional focus on parent-child concordance to advocate for interventions that involve the full household, and not only prevention for adults.

In the present study, we analyzed the concordance of maternal and child weight status, a critical risk factor for NCDs. We further investigate the dynamics of these associations across different age groups of children under 5.

## Materials and Methods

### Data sources and participants

Pakistan’s Demographic and Health Survey, Round VII (PDHS-7) survey was conducted in 2017-18 by the National Institute of Population Studies and funded by the United States Agency for International Development (USAID). PDHS-7 was the 7^th^ round of a nationally representative survey, with information about household, demographic, and maternal and child health indicators. The survey design and sample size calculations were formulated to provide reliable information at the national level and subnational levels, as well as for urban and rural areas separately, with data collected from the four provinces of Punjab, Sindh, Khyber Pakhtunkhwa, and Balochistan, the regions of Azad Jammu and Kashmir and Gilgit Baltistan, the Islamabad Capital Territory, and the Federally Administered Tribal Areas. The sampling design used a two-stage stratified approach and, because of non-proportional sample allocation, the sample weights were generated by the PDHS team. Participants were recruited and data was collected from November 2017 to April 2018

The respondents included all ever-married women aged 15-49 in all selected households. In one-third of the selected households, height and weight were directly measured for ever-married women 15-49 and children under-five years of age. For this study, we included all children under-five who had WHZ information and BMI information on their mothers, leading to a sampling-weight adjusted sample size of 3465. Fig 1 depicts the sampling scheme and relevant response rates for PDHS-7 [21].

**Fig 1.**
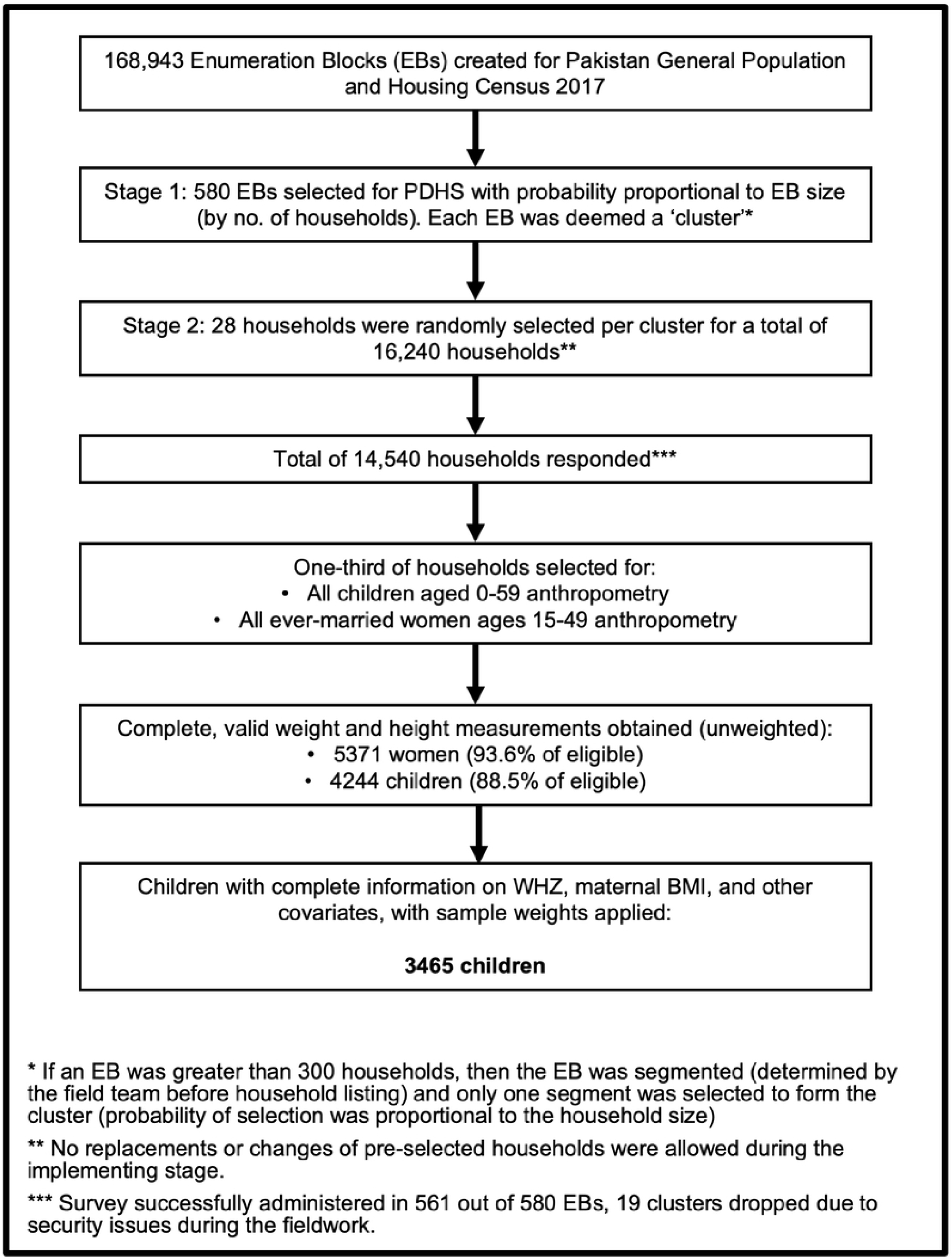
Flow diagram of household and participant selection for the 2017-18 Pakistan Demographic and Health Survey.

The study was reviewed and approved by the National Bioethics Committee of the Pakistan Health Research Council and the Inner City Fund/ ICF Institutional Review Board. Adult participants were consented both, for themselves and for their children [21]. Prior to obtaining access to and performing analysis on the PDHS-7 data, the researchers of the present study submitted a concept note, gained data access, and obtained permission from USAID. The data were de-identified and anonymized by the DHS program prior to being shared, therefore not considered human subjects research by an author-affiliated institution (Emory University, Atlanta, GA, USA) and not requiring further ethical clearance.

### Study Measures

#### Child weight status: Weight-for-height z-score (WHZ)

Child weight and height measurements were taken during a single visit. Weight measurements were taken using SECA scales (model no. SECA 878U) and height measurements using a Shorr Board (recumbent if child’s age was less than 24 months and standing if older). Anthropometry data in PDHS-7 was collected by two female enumerators in each field team (total of 44 enumerators) who jointly took measurements. All enumerators were trained beforehand to standardize procedures for anthropometry, including hands-on training to measure ten children twice to assess accuracy and precision of measurements and further training for those enumerators who were out of range more three or more times [21].

The dependent variable was weight-for-height z-score (WHZ) for children under-five per the 2006 Child Growth Standards released by the World Health Organization, reliably shown to describe both overweight (WHZ > 2) and wasting (WHZ < -2) [22]. Every one unit in z-score represents one standard deviation from the mean weight-for-height from a study of more than eight thousand children recruited from Brazil, Ghana, India, Norway, Oman and the United States of America [23,24]. For example, a WHZ of -2 indicates that the child’s weight is two standard deviations below the mean for their height and sex [25].

#### Maternal weight status: Body Mass Index (BMI)

The primary exposure of interest for this study was maternal BMI. BMI was measured in the same way as child WHZ, with SECA 878U scales and Shorr Boards (standing height measurement). Maternal BMI was coded as obese (30 or greater), overweight (25 to less than 30), healthy weight (18.5 to less than 25), and underweight (less than 18.5), which was the reference category ([ref]).

##### Covariates

Several other covariates were also considered in the analysis. A covariate was included in statistical the models if it had evidence of correlation with child weight outcome and if there was sufficient variation in the covariate for it to add information to the model, e.g. if a factor has 99% ‘yes’ response rate and 1% ‘no’ rate, then its use was avoided because it would not meaningfully explain variation in the outcome. Through this process, the following eight covariates were selected for analysis: urban/ rural specific wealth index [18,22,26–28], sex of child [22,26], mother’s employment status[26,27,29], type of place of residence [28,30–32], mother’s youngest child under-five [28,33,34], mother’s oldest child under-five [28,33,34], child age in months, and maternal age in years [29,33,34].

Household wealth status was measured as quintiles of assets as provided by the PDHS-7 survey. Wealth index quintile categories were labeled as richest, rich, middle, poor, poorest[ref]. Child sex was coded as male or female[ref]. Type of place of residence was coded as urban or rural[ref]. Mother’s employment status, youngest child under-five and oldest child under-five were all codes as no or yes[ref].

### Analysis

A dyadic dataset of mothers and their children was created by merging anthropometric and demographic characteristics of mothers and children per DHS guidance for analysis. Mother’s characteristics were treated as exposures for the child [35]. Furthermore, in studying cardiometabolic risk concordance between younger children and their mothers, it is important to consider the impact of children being born small for gestational age (SGA) can have on anthropometric concordance. This is particularly important consideration in Pakistan, where the prevalence of SGA births is >45% [36]. As approximately 85% of children born SGA have experienced sufficient catch up growth by age 2 [37–39], stratifying analysis by this age point in relation to concordance which, to our knowledge, has not been done before, could yield important findings regarding the ‘unmasking’ of parent-child concordance. Thus, we stratified children by two years of age in our analysis and modeling.

All analysis and modeling accounted for the complex survey design [35]. All data restructuring and analysis was done using IBM SPSS Statistics v29.0.0.0 with Complex Samples v29. The code used for restructuring and analysis can be found via the following link: doi:10.5281/zenodo.7794384.

#### Descriptive statistics

Reported statistics (sampling weight-adjusted) included background characteristics of the participants which include average age of mother (in years) and children (in months), mean BMI of mothers, and mean child WHZ with 95% confidence intervals. Other reported adjusted population descriptive statistics were percentage of dyads that were urban vs rural, percentage of male and female children, percentage of mothers currently employed, percentage of children being the youngest child, percentage of children being the oldest under-five, and the percentage of the sample size in each wealth index quintile. The mean child WHZ with 95% confidence interval and sampling weight-adjusted sample size was also reported for each factor level.

#### Modeling associations between maternal and child weight status

We estimated linear regression models to investigate concordance between maternal BMI and child WHZ, measured continuously. Three linear models were generated: 1) all children under-five years, 2) all children under-two years, 3) all children ages 2 to 5, with the models adjusted for the 8 covariates. We reported the R^2^ value and the Wald F for each model and reported the parameter estimates with 95% confidence interval and t-test value for each categorical level. For each model, the statistically significant variables were remodeled including only these variables and an additional term for each two-variable combination to assess for interaction between the statistically significant variables. Cases with missing values for any variables were treated as invalid. Associations for all models were considered statistically significant when p < 0.05.

#### Further Considerations for Analysis

We assessed multicollinearity between the independent variables using the variance inflation factor (VIF) calculation. We used a conservative cutoff of 3, as an indicator to further investigate relationships [40]. Using this cutoff, no multicollinearity was observed among the independent variables. To account for multiple comparisons and counter the increased risk of Type I error, the Holm-Bonferroni method was used to adjust the P-value of 0.05 accordingly [41,42]. Regarding outliers, the DHS data is screened before dissemination for plausibility of Z-scores. For children who had WHZ that were below -5 standard deviations or above +5 standard deviations were flagged and their WHZ was not reported. Similarly, women who had BMI below 12 or above 60 were also flagged and did not have their BMI reported [35]. Further consideration of outliers was not made because of these adjustments by the PDHS data processing team.

## Results

For the analysis, out of 4671 sample unweighted cases, 4130 had complete variable information (with 541 missing) and were included, which led to a sampling-weight adjusted sample size of 3465. With cases that had information on maternal BMI and child WHZ, only two did not have complete information on the other covariates.

### Summary Statistics

Of the overall sampling weight-adjusted sample of 3465 children (Fig. 1), 1406 (40.6%) children were less than 2 years old, and 2059 (59.4%) children were ages 2 to 5. The mean age of mothers was 29.0 years (95% CI [28.6, 29.4]) and children under-five years was 28.8 months (95% CI [28.0, 29.5]). The mean maternal BMI was 24.6 (95% CI [24.2, 25.1]). The mean child WHZ for the entire sample was -0.30 (95% CI [-0.36, -0.24]), mean child WHZ for children under-two was -0.41 (95% CI [-0.50, -0.31]), and mean child WHZ for children ages 2 to 5 was -0.23 (95% CI [-0.29, -0.16]). The greatest mean child WHZ was in children who belonged to the richest quintile of households at 0.04 (95% CI [-0.09, 0.17]), and the category with the lowest mean child WHZ was children with mothers who were underweight at -0.66 (95% CI [-0.80, -0.51]). Table 1 summarizes maternal, child, and household characteristics of the analyzed maternal-child dyads and the mean child WHZ for each factor level is summarized in Table 2.

**Table 1:** Percentage of sample in each factor level and mean values of continuous variables (n=3465), with 95% confidence interval from PDHS-7 dataset used in analysis.

| Variable | All | Children under two | Children 2 to 5 years |
| --- | --- | --- | --- |
| **Maternal Characteristics** |  |  |  |
|  | **Percent (95% CI)** |  |  |
| **Maternal BMI Category** |  |  |  |
| 30+ | 15.7 (13.1 – 18.8) | 13.1 (10.4 – 16.4) | 17.5 (14.5 – 20.9) |
| 25 to < 30 | 27.6 (24.8 – 30.6) | 27.9 (24.5 – 31.7) | 27.4 (24.3 – 30.7) |
| 18.5 to <25 | 46.2 (42.9 – 49.6) | 46.7 (42.6 – 50.8) | 45.9 (42.1 – 49.8) |
| <18.5 | 10.4 (8.5 – 12.7) | 12.3 (9.9 – 15.1) | 9.2 (7.2 – 11.7) |
| **Mother currently employed** |  |  |  |
| No | 86.7 (83.9 – 89.0) | 34.6 (30.1 – 39.4) | 31.4 (27.6 – 35.5) |
| Yes | 13.3 (11.0 – 16.1) | 65.4 (60.6 – 69.9) | 68.6 (64.5 – 72.4) |
|  | **Mean (95% CI)** |  |  |
| **Maternal BMI** | 24.6 (24.2, 25.1) | 24.3 (23.8, 24.8) | 24.9 (24.4, 25.4) |
| **Maternal Age (yrs.)** | 29.0 (28.6, 29.4) | 27.4 (26.9, 27.9) | 30.3 (29.6, 30.5) |
| **Child Characteristics** |  |  |  |
|  | **Percent (95% CI)** |  |  |
| **Youngest child** |  |  |  |
| No | 34.3 (32.8 – 35.9) | 5.5 (4.2 – 7.2) | 54 (51.3 – 56.7) |
| Yes | 65.7 (64.1 – 67.2) | 94.5 (92.8 – 95.8) | 46 (43.3 – 48.7) |
| **Oldest child under-five** |  |  |  |
| No | 32.5 (30.9 – 34.1) | 56.4 (52.9 – 59.8) | 16.2 (14.3 – 18.2) |
| Yes | 67.5 (65.9 – 69.1) | 43.6 (40.2 – 47.1) | 83.8 (81.8 – 85.7) |
| **Sex of child** |  |  |  |
| Male | 51.1 (49.1 – 53.1) | 51.2 (47.3 – 55.0) | 51.0 (48.7 – 53.3) |
| Female | 48.9 (46.9 – 50.9) | 48.8 (45.0 – 52.7) | 49.0 (46.7 – 51.3) |
|  | **Mean (95% CI)** |  |  |
| **Child WHZ** | -0.30 (-0.36, -0.24) | -0.41 (-0.50, -0.31) | -0.23 (-0.29, -0.16) |
| **Child Age (mos.)** | 28.8 (28.0, 29.5) | 11.0 (10.5, 11.5) | 40.9 (40.4, 41.5) |
| **Household Characteristics** |  |  |  |
|  | **Percent (95% CI)** |  |  |
| **Area** |  |  |  |
| Urban | 32.7 (29.1 – 36.6) | 34.6 (30.1 – 39.4) | 31.4 (27.6 – 35.5) |
| Rural | 67.3 (63.4 – 70.9) | 65.4 (60.6 – 69.9) | 68.6 (65.4 – 72.4) |
| **Wealth index (urban/ rural specific)** |  |  |  |
| Richest | 18.3 (14.7 – 22.5) | 19.1 (14.7 – 24.5) | 17.7 (14.1 – 21.9) |
| Richer | 20.3 (17.0 – 24.0) | 21.9 (18.0 – 26.3) | 19.1 (15.7 – 23.1) |
| Middle | 20.1 (17.4 – 23.0) | 20.2 (17.0 – 23.9) | 19.9 (17.0 – 23.2) |
| Poor | 21.4 (18.4 – 24.8) | 20.2 (16.8 – 24.1) | 22.3 (18.8 – 26.1) |
| Poorest | 20.0 (16.5 – 24.0) | 18.6 (15.1 – 22.5) | 21.0 (16.8 – 25.8) |

**Table 2:** Mean Child WHZ and sampling weight-adjusted sample size (Adj. N) of analyzed sample, stratified by level of selected factors.

| Variable | Adj. N | Mean child WHZ (95% CI) |
| --- | --- | --- |
| **All** | 3465 | -0.3 (-0.36, -0.24) |
| **Maternal BMI Category** |  |  |
| 30+ | 545 | -0.02 (-0.16, 0.11) |
| 25 to < 30 | 956 | -0.14 (-0.25, -0.03) |
| 18.5 to <25 | 1602 | -0.41 (-0.51, -0.31) |
| <18.5 | 361 | -0.66 (-0.8, -0.51) |
| **Mother currently employed** |  |  |
| No | 3003 | -0.29 (-0.36, -0.23) |
| Yes | 462 | -0.35 (-0.52, -0.19) |
| **Youngest child** |  |  |
| No | 1189 | -0.21 (-0.29, -0.13) |
| Yes | 2276 | -0.35 (-0.42, -0.28) |
| **Oldest child under-five** |  |  |
| No | 1126 | -0.4 (-0.51, -0.28) |
| Yes | 2339 | -0.26 (-0.32, -0.2) |
| **Sex of child** |  |  |
| Male | 1770 | -0.32 (-0.39, -0.24) |
| Female | 1695 | -0.29 (-0.36, -0.21) |
| **Area** |  |  |
| Urban | 1134 | -0.31 (-0.4, -0.21) |
| Rural | 2331 | -0.30 (-0.37, -0.22) |
| **Wealth index (urban/ rural specific)** |  |  |
| Richest | 633 | 0.04 (-0.09, 0.17) |
| Richer | 702 | -0.17 (-0.29, -0.06) |
| Middle | 695 | -0.36 (-0.5, -0.22) |
| Poor | 743 | -0.52 (-0.64, -0.4) |
| Poorest | 693 | -0.45 (-0.58, -0.31) |

### Associations between mother and child weight status

Maternal BMI was positively associated with child WHZ for all children among children under-five. Children of normal weight, overweight, and obese women had WHZ scores that were 0.22 [95% CI: 0.05, 0.38], 0.44 [95% CI: 0.26, 0.63], and 0.53 [95% CI: 0.33, 0.73] units higher than children of underweight women, respectively. Maternal BMI was also positively associated with child WHZ among children ages 2 to 5, where children of normal weight, overweight, and obese women had WHZ scores that were 0.27 [95% CI: 0.09, 0.44), 0.51 [95% CI: 0.30, 0.72), and 0.61 [95% CI: 0.37, 0.85] units higher than children of underweight women, respectively.

For all children under-five, household wealth index was positively associated with child WHZ (Richest: 0.37 [95% CI: 0.18, 0.55]; Rich: 0.19 [95% CI: 0.02, 0.37]; Middle: 0.01 [95% CI: -0.17, 0.20]; Poor: -0.12 [95% CI: -0.30, 0.06]; Poorest [ref.]). Household wealth was similarly positively associated with child WHZ for children under 2 (Richest: 0.44 [95% CI: 0.13, 0.74]; Rich: 0.21 [95% CI: -0.08, 0.49]; Middle -0.31 [95% CI: -0.58, - 0.44]; Poor: -0.15 [95% CI: -0.46, 0.17]; Poorest: [ref]) and for children aged two to five (Richest: 0.31 [95% CI: 0.11, 0.51]; Rich: 0.17 [95% CI: -0.01, 0.35]; Middle 0.23 [95% CI: 0.03, 0.43]; Poor -0.10 [95% CI: -0.28, 0.07]; Poorest: [ref.]).

There were no statistically significant interaction effects for any linear model. Table 3 summarizes information for each model.

**Table 3:** Linear regression results of the association between maternal BMI and child WHZ, along with selected covariates, among three population groups: all children under-five, children under-two, children 2 to 5.

| Variable<br>(Category) | All children under-five |  |  | Children under-two |  |  | Children 2 to 5 years |  |  |
| --- | --- | --- | --- | --- | --- | --- | --- | --- | --- |
|  | Coefficient (95% CI) | t-value | p | Coefficient (95% CI) | t-value | p | Coefficient (95% CI) | t-value | p |
| **Mother BMI category** |  |  | **(<0.001)** |  |  | **0.130** |  |  | **(<0.001)** |
| 30 or greater | 0.53(0.33, 0.73) | 5.201 | <.001 | 0.32(-0.02, 0.67) | 1.836 | 0.067 | 0.61(0.37, 0.85) | 5.043 | <.001 |
| 25 to <30 | 0.44(0.26, 0.63) | 4.670 | <.001 | 0.26(-0.05, 0.57) | 1.642 | 0.101 | 0.51(0.30, 0.72) | 4.857 | <.001 |
| 18.5 to <25 | 0.22(0.05, 0.38) | 2.587 | 0.010 | 0.06(-0.21, 0.34) | 0.447 | 0.655 | 0.27(0.09, 0.44) | 2.945 | 0.003 |
| less than 18.5 | * |  |  | * |  |  | * |  |  |
| **Mother currently working** |  |  | **0.363** |  |  | **0.812** |  |  | **0.168** |
| No | -0.08(-0.25, 0.09) | -0.910 | 0.363 | 0.03(-0.21, 0.28) | 0.265 | 0.791 | -0.14(-0.33, 0.05) | -1.423 | 0.155 |
| Yes | * |  |  | * |  |  | * |  |  |
| **Youngest child** |  |  | **0.020** |  |  | **0.024** |  |  | **0.065** |
| No | 0.15(0.02, 0.27) | 2.335 | 0.020 | 0.36(0.05, 0.67) | 2.285 | 0.023 | 0.11(-0.03, 0.25) | 1.574 | 0.116 |
| Yes | * |  |  | * |  |  | * |  |  |
| **Oldest child under-five** |  |  | **0.519** |  |  | **0.650** |  |  | **0.667** |
| No | -0.05(-0.19, 0.10) | -0.646 | 0.519 | -0.05(-0.24, 0.14) | -0.491 | 0.624 | -0.05(-0.25, 0.16) | -0.445 | 0.657 |
| Yes | * |  |  | * |  |  | * |  |  |
| **Sex of child** |  |  | **0.420** |  |  | **0.178** |  |  | **0.934** |
| Male | -0.04(-0.14, 0.06) | -0.808 | 0.420 | -0.12(-0.3, 0.06) | -1.358 | 0.175 | 0.00(-0.10, 0.11) | 0.045 | 0.964 |
| Female | * |  |  | * |  |  | * |  |  |
| **Area** |  |  | **0.474** |  |  | **0.768** |  |  | **0.455** |
| Urban | -0.04(-0.15, 0.07) | -0.723 | 0.470 | -0.03(-0.21, 0.16) | -0.287 | 0.774 | -0.05(-0.19, 0.08) | -0.785 | 0.433 |
| Rural | * |  |  | * |  |  | * |  |  |
| **Wealth index (urban/ rural specific)** |  |  | **(<.001)** |  |  | **(<.001)** |  |  | **(<.001)** |
| Richest | 0.37(0.18, 0.55) | 3.942 | <.001 | 0.44(0.13, 0.74) | 2.825 | 0.005 | 0.31(0.11, 0.51) | 3.088 | 0.002 |
| Rich | 0.19(0.02, 0.37) | 2.155 | 0.032 | 0.21(-0.08, 0.49) | 1.413 | 0.158 | 0.17(-0.01, 0.35) | 1.812 | 0.071 |
| Middle | 0.01(-0.17, 0.2) | 0.139 | 0.889 | -0.31(-0.58, -0.04) | -2.271 | 0.024 | 0.23(0.03, 0.43) | 2.227 | 0.026 |
| Poor | -0.12(-0.3, 0.06) | -1.341 | 0.181 | -0.15(-0.46, 0.17) | -0.916 | 0.360 | -0.10(-0.28, 0.07) | -1.168 | 0.244 |
| Poorest | * |  |  | * |  |  | * |  |  |
| **Mother age (yrs)** | 0.00(-0.01, 0.01) | -0.130 | 0.897 | 0.00(-0.02, 0.02) | 0.229 | 0.819 | -0.01(-0.02, 0.00) | -2.490 | **0.019** |
| **Child age (mos)** | 0.00(0.00, 0.01) | 0.340 | 0.734 | 0.01(0.00, 0.02) | 1.312 | 0.190 | -0.01(-0.02, 0.00) | -2.490 | **0.528** |
| **R<sup>2</sup>** | **0.050** |  |  | **0.060** |  |  | **0.064** |  |  |
| **Wald F** | **8.551** |  |  | **3.885** |  |  | **9.147** |  |  |

Overall variable p-values in parenthesis denote statistically significant associations (Holm-Bonferroni corrected)
\* denotes reference category

## Discussion and Conclusion

We found that maternal BMI was positively associated with child WHZ in children under 5. This positive association was present for children ages 2 to 5, even after accounting for several socio-demographic factors, but not for children under-two. The findings suggest that there is intergenerational concordance in weight status in Pakistani mothers and young children which is unmasked with increased age.

The findings of parent-child concordance of BMI-WHZ for all children under-five are consistent with previous literature which specifically explore parent-child concordance of NCD risk [2,11–16] and of association studies in South Asia which have observed a positive association between maternal and children under-five weight status [17,18]. However, previously described phenomenon of the double burden of malnutrition at household level (DBMHL) suggests co-existence of undernourished children and overweight mothers in the same household, but this phenomenon is primarily described for child stunting and, in some cases, underweight [43–51]. Although a recent study by Biswas et al., in which they analyzed the co-existence of overweight mothers with all types of undernourished children under-five (stunted, wasted, and underweight) in South and Southeast Asia from 2006 to 2017 data, described positive relationships between maternal weight status and all three types of child undernutrition, with the greatest co-existence in Pakistan. However, this study did not analyze the relationship between non-overweight or obese mothers and their children, nor did it split the children under-five into two age groups [52].

Furthermore, double burden of malnutrition (DBM) at the country or population level is well-established in previous literature [53–56]. DBM prevalence has been increasing in Pakistan as shown by PNNS 2018 results, with wasting noted in 17.7% of children, the highest in Pakistan’s history, prevalence of overweight children under-five almost doubling from 5% in 2011 to 9.5% in 2018, and women ages 15-49 experiencing an increase in overweight and obesity increasing from 28% to 37.8% from 2011 to 2018. Both, DBMHL and DBM, can co-exist with maternal-child BMI-WHZ concordance, as the increasing burden of DBM, with an increasing percentage of overweight children, could be leading to the more recent findings of positives association of maternal-child weight status. Co-existence of these phenomena within the same society highlight the complexity of interventions and approaches required to address malnutrition among LMICs, as different regions and households are experiencing various challenges.

To our knowledge, there are no studies which specifically investigate the relationship between maternal and child concordance under-five stratified by 2 years of age, as we analyzed in this study. These findings, which show a positive association between maternal BMI and child WHZ only in the 2 to 5 age group, potentially indicate that concordance between child WHZ and maternal BMI is ‘unmasked’ after age 2, when the majority of early childhood catch-up growth has occurred [37–39]. As in, children who may be born SGA may not show concordance of WHZ with maternal BMI during the first two years of life, but as they experience their expected catch up growth and have spent more time in a shared environment with their mothers, their weight status starts to become concordant with their mothers, as seen by our analysis which shows the difference in association between children older than and younger than two. This suggests that children under-two who do not initially appear to have WHZ concordant to their mother’s BMI may develop this positive relationship later in life. Furthermore, a targeted review on maternal and child overweight and obesity in LMICs shows that junk food, such as potato chips, sponge cakes, sugary biscuits, and sugary drinks are a significant part of child diet by the first two years of life [57], potentially yielding an avenue of intervention and an explanation for the development of mother-child concordance after two years of age. This relationship could also be affected by other dietary aspects such as breastfeeding status and weaning, as prolongation of breastfeeding has been shown to decrease excess weight gain in children [58]. Overall, this may indicate an avenue for early interventions for weight-related risks in children, where healthier nutritional sources during infancy may lead to a decreased sharing of NCD risk found between very young children and their mothers as children age.

Regarding other covariates which were analyzed, we showed wealth index as a statistically significant positive factor across all GLM analyses. This association existing in Pakistan is well established in the literature [59–64]. Increasing wealth quintile has been suggested to correlate with access to nutrient rich foods and increased sedentary behaviors [18,65–68].

Considering strengths of this study, the analysis was done on a nationally representative dataset which involved a complex sampling process to obtain the best possible demographic snapshot of Pakistan. The analysis also included a significant number of covariates to unveil potential confounders and used rigid levels of correction and validation regarding significance and multi-collinearity. Limitations of the study included the use of cross-sectional data, whereas a cohort study could provide more in-depth data on trends in specific households.

Overall, the findings of maternal-child concordance and, more specifically, the ‘masking’ phenomenon could have important implications regarding interventions and policy making, where these seemingly non-concordant groups may be ignored as an avenue to improve NCD risk unless there is data suggesting that the concordance may ‘unmask’ later. Future work should involve more detailed data collection of cardiometabolic disease and, more broadly, NCD risk concordance and outcome concordance on entire households, including young children, to develop family-wide interventions to reduce long-term mortality from these diseases.

## Data Availability

The PDHS dataset is available at: https://dhsprogram.com/. Prior to obtaining access to and performing analysis on the PDHS-7 data, a concept note may be required, as was the case for the authors of the manuscript. The code used for data restructuring and analysis is available at: doi:10.5281/zenodo.7794384

doi:10.5281/zenodo.7794384

https://dhsprogram.com/

## Acknowledgements

We thank the DHS Program supported by USAID, the National Institute of Population Studies, and ICF for the data used in this study. We also thank the Emory Global Diabetes Research Center and Global Health Equity Scholars NIH FIC TW010540.

